# Effect of motivational interviewing on lifestyle modification among patients with hypertension attending the family medicine clinics of Irrua Specialist Teaching Hospital, Irrua, Nigeria – A randomized controlled trial (MILMAPH Study)

**DOI:** 10.1101/2024.04.16.24305912

**Authors:** Tijani Idris Ahmad Oseni, Afiong Oboko Oku, Ifeoma N Monye, Sulaiman Dazumi Ahmed, Roseline Ekanem Duke, Osahon Otaigbe, Ndifreke Ekpo Udonwa

**Affiliations:** Department of Family Medicine, Edo State University, Uzairue, Edo State, Nigeria; Department of Community Medicine, University of Calabar, Calabar, Nigeria; Brookfield Clinics Centre for Lifestyle Medicine, Abuja, Nigeria; Department of Internal Medicine, Irrua Specialist Teaching Hospital, Irrua, Nigeria; Faculty of Clinical Sciences, University of Calabar, Calabar, Nigeria; Department of Community Medicine, Irrua Specialist Teaching Hospital, Irrua, Nigeria; Department of Family Medicine, University of Calabar, Calabar, Nigeria

**Keywords:** motivational interviewing, hypertension, blood pressure control, lifestyle modification, Nigeria

## Abstract

**Background:** Lifestyle modification has been shown to improve blood pressure (BP) control, but its practice is poor among patients with hypertension. Thus, measures that would help patients with hypertension achieve positive lifestyle modification would improve BP control. The study aimed to determine the effect of motivational interviewing on lifestyle modification and blood pressure control among patients with hypertension attending the Family Medicine Clinics of Irrua Specialist Teaching Hospital (ISTH), Irrua, Nigeria.

**Methods:** The study was a randomised control trial (PACTR202301917477205) of 250 hypertensive adults between 18 and 65 years presenting to the Family Medicine Clinics of ISTH randomised into intervention and control groups. Those in the intervention group were given monthly motivational interviewing (MI) on lifestyle modification in addition to standard care for the management of hypertension while those in the control group got standard care only for 6 months. Both groups were assessed at baseline and 6 months. Data was analysed with Stata version 17 (StataCorp LLC) with level of significance at 0.05. Primary outcome was lifestyle modification while secondary outcome was BP control both at 6 months.

**Results:** The age of the participants ranged from 24 to 65 years with a mean age of 51.5 ± 10.0 years. The blood pressure control at baseline was 24%. At the end of the six-month study, there was a statistically significant improvement in the lifestyle pattern of the intervention group compared to control except smoking in which the difference was not statistically significant (p=0.150). Blood pressure control significantly improved from 24% at baseline to 48% post-intervention (p=0.014).

**Conclusion:** The study found a significant positive association between motivational interviewing and lifestyle modification. The study also found a significant improvement in blood pressure control following motivational interviewing. There is a need to incorporate motivational interviewing into the management of hypertension for better outcomes.

## Introduction

Hypertension is a leading cause of morbidity and mortality globally.^1^ It is the leading cause of preventable death globally.^2^ It is also the leading global disease burden accounting for over 80%.^3,4^ According to the World Health Organization, an estimated 1.28 billion adults aged 30 to 79 years globally are hypertensive with two-thirds of them in low- and middle-income countries.^5^ Hypertension is the leading risk factor by attributable disability-adjusted life-years (DALYs).^6–8^ It accounts for 15 million deaths of persons 30 to 69 years with 80% of these premature deaths occurring in low- and medium-income countries including Nigeria.^9^

Risk factors for hypertension include older age, genetic predisposition, being overweight or obese, not being physically active, high-salt diet, poor sleep, inadequate fruit and vegetable intake, smoking and excess alcohol consumption.^5,6,10–13^

Blood pressure control among hypertensives is poor both globally and nationally.^14–16^ According to the World Health Organisation (WHO), only 21% of adults with hypertension have their blood pressure controlled.^5^ One in four patients with hypertension is unable to achieve adequate blood pressure control despite medications.^17–19^ The limited health insurance coverage in Nigeria resulting in out-of-pocket spending leads to catastrophic health expenditure and further worsens the already poor blood pressure control among Nigerian hypertensives.^4^

Lifestyle modification is key to the prevention and control of hypertension as well as the reduction in the risk of CVD and is therefore recommended in all major guidelines for the management of hypertension.^20–24^ It has also been advocated by the WHO as the first-line treatment for uncomplicated hypertension with drug therapy only initiated if control is not achieved with lifestyle modification alone.^5^ Lifestyle modification is also advocated to be continued even after initiating drug therapy, as it has been shown to improve outcomes and reduce the risk of CVD.^5,20–24^ Healthy lifestyle practices like increased physical activity, healthy diet, dietary salt reduction, reduction/cessation of alcohol consumption, smoking cessation and weight control, reduced blood pressure and improved control of hypertension and cardiovascular health.^2,6,25^ However, adherence to recommended lifestyle modification is very low despite routine lifestyle modification counselling.^26–30^

Motivational interviewing (MI) is a focused patient-centred and goal-directed counselling technique that motivates individuals to change by exploring and resolving ambivalence.^31^ The counsellor evokes the client to make his/her own decision to change by facilitating the process of change.^32^ This is important as studies have shown that hypertension can be controlled and this control is mainly by the patient.^33–35^ Motivational interviewing helps patients with hypertension achieve control if properly implemented. It is a special type of patient-centred counselling that leverages on the framework that accesses risk behaviour, advises change, agrees on goals and plans through shared decision-making, assists with treatment and arranges for follow-up.^34–38^ It has been used to effect changes in health behaviour through self-reflection thereby helping individuals prevent unhealthy behaviours and promoting healthy behaviours.^35,37^ This has resulted in weight loss in overweight or obese patients and, a reduction in triglyceride levels, waist circumference, and systolic blood pressure.^34^

This study aimed to determine the effect of Motivational Interviewing on lifestyle modification and blood pressure control among patients with hypertension in the Family Medicine Clinics of Irrua Specialist Teaching Hospital, Irrua, Edo State, South-South Nigeria.

### Conceptual Framework

This study used the transtheoretical model of change that was developed by Proschaska and Diclemente to describe individual behaviour change in dealing with addictions.^39^ The study was built on the concept that MI would help motivate patients with hypertension to adopt desired healthy lifestyle habits of physical activity, salt reduction, fruit and vegetable intake, smoking cessation and alcohol reduction/cessation. This would in turn lead to blood pressure control as established by studies. The conceptual framework is illustrated in Fig 1.

**Fig 1:**
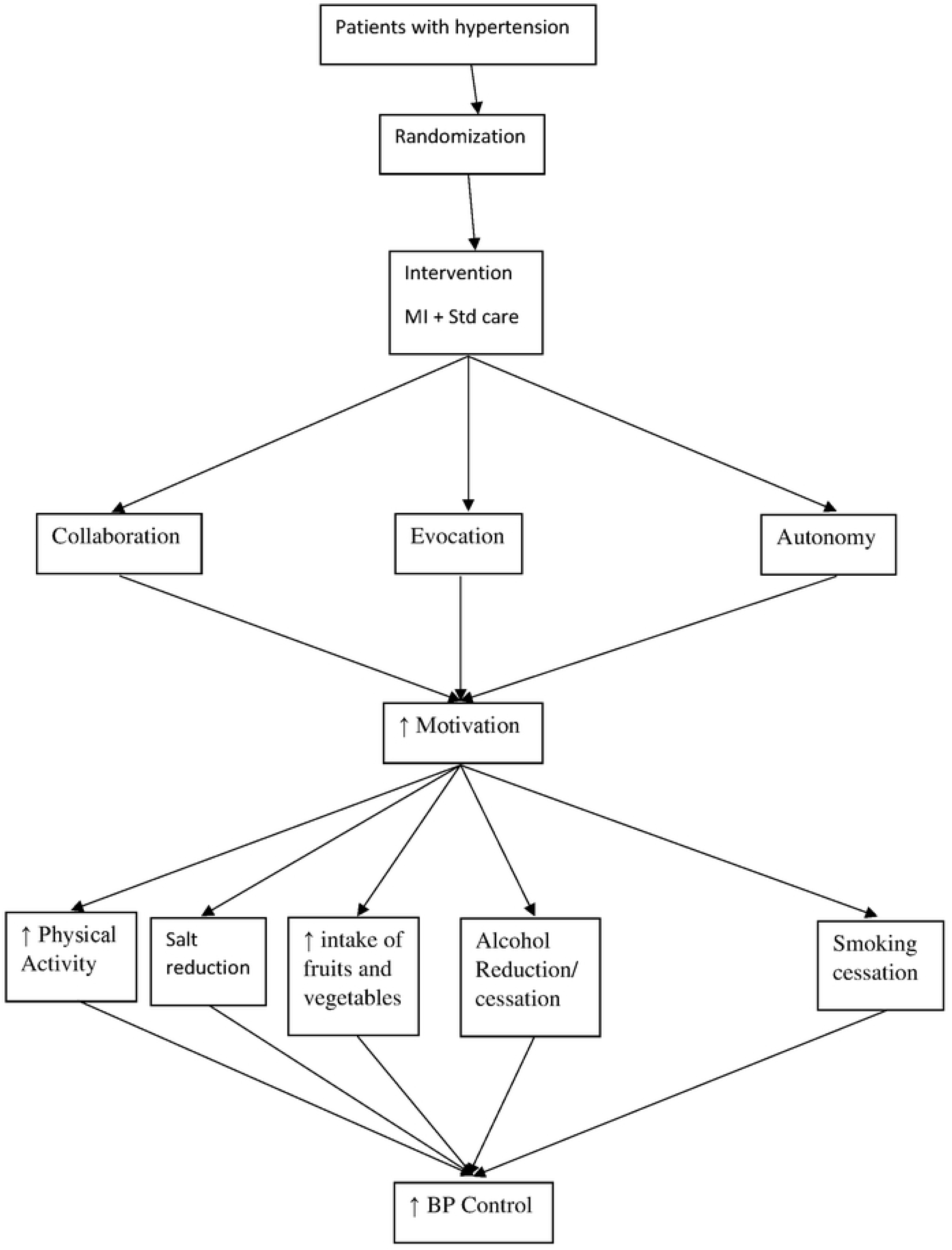
Conceptual Framework

## Methods

### Study Design

The study was a double-blind, parallel-group randomised controlled trial (RCT). It was registered with the Pan African Clinical Trial Registry with trial registration no PACTR202301917477205. The protocol was designed using the SPIRIT 2013 Checklist for clinical trial protocol and related documents and published in the West African Journal of Medicine.^40^ The study was conducted between 17^th^ April 2023 and 1^st^ December 2023 and reported using the CONSORT 2010 checklist for reporting a randomised trial.

### Study Setting

The study was conducted in the Family Medicine Clinics of Irrua Specialist Teaching Hospital (ISTH), a tertiary hospital in Irrua, a semi-urban town and headquarters of Esan Central Local Government Area in Edo Central Senatorial District, Edo State, South-South Nigeria. Irrua has an area of 4,346 KM^2^ and a population of 60,545 with 50.4 per cent of them being males.^41^ The Family Medicine Clinics consist of the General Outpatient Department (GOPD) and the Staff/National Health Insurance Authority (NHIA) clinics. All patients presenting to the hospital are first seen in the Family Medicine Clinics except for emergencies. Hospital records show that between 70 to 80 per cent of patients with hypertension attending ISTH are seen at the Family Medicine clinics. The clinics see about 400 patients with hypertension monthly with about 300 seen in the GOPD and the remaining 100 seen in Staff/NHIA clinic respectively.

### Study Population

Adult patients aged 18 to 65 years with hypertension, including newly diagnosed patients, presenting to the Family Medicine Clinics of the hospital who consented to the study. Exclusion criteria included Patients with cognitive impairment (assessed through a mini-mental state examination conducted for all participants at baseline) who may not be able to understand or comprehend well; those with physical disability that would limit them from exercising; the severely ill patients whose conditions would not allow them to be interviewed on review dates; those with hypertensive complications like renal disease and stroke (Renal disease was ruled out through a baseline urinalysis test for all patients and patients with marked proteinuria were excluded from the study); as well as pregnant women (A pregnancy test was done for all female patients of reproductive age and those who tested positive were excluded from the study).

### Sample Size Estimation

The sample size was determined at a confidence level of 95 per cent using the sample size formula for RCT by Walters:^42^

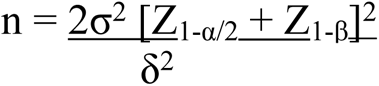

where n is the sample size per group; σ = Standard Deviation; Z_1-α/2_ is the Z value corresponding to 95% level of significance = 1.96, Z_1-β_ is the Z value corresponding to 80 per cent power = 0.84 and δ is the target or anticipated difference in mean outcomes between the two groups. Steffen et al^43^ in a study to evaluate the effectiveness of motivational interviewing in the management of Type 2 diabetes and arterial hypertension in primary healthcare in Brazil found the mean reduction in systolic BP to be 14.4 ± 10.8. Using the standard deviation from Steffen et al, the sample size was calculated as follows:

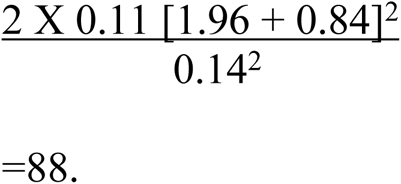

Assuming an attrition rate of 20 percent, (which approximates to 18), the minimum sample size was calculated to give 106. Therefore, 106 patients were to be recruited for each arm of the study, bringing the total number of participants to 212. However, 125 patients were recruited in each arm of the study to increase the power of the study.

### Recruitment and Randomization

Adults with hypertension presenting to the Family Medicine Clinics were recruited using systematic sampling. The Family Medicine clinics of ISTH attend to about 400 patients with hypertension monthly. The sampling interval was calculated by dividing the population of the study (400 patients monthly for the 3 months (April to June 2023) the baseline data collection lasted=1200) by the calculated sample size=1200/303=3.96≈4. A sampling interval of 1:4 was used as it gave all eligible subjects equal chances of being selected within the period of study. The first patient was selected from the first four patients with hypertension by simple random sampling; subsequently, every fourth patient was selected. However, if the prospective participant to be selected was not eligible or refused to participate, the next eligible participant was selected. Each patient’s card was tagged. The identification sticker was left on all selected cards until the study was over to avoid a repeat selection. This was done on every clinic day (Mondays to Fridays except on public holidays) until the sample size was achieved. Proportionate recruitment of patients from both the GOPD and the Staff/NHIA clinic was done at a 3:1 ratio as hospital records showed that the GOPD saw about three times the number of patients with hypertension than the Staff/NHIA clinic.

Patients were then randomised into Intervention and Control Groups. Blinding of the participants, data collectors, and analysts was ensured throughout the process. During randomisation, opaque envelopes were numbered serially and with cards to indicate if a patient belonged to either the intervention or control arm. Randomisation was done by professionals who were not part of the research team. The allocation ratio was 1:1. Blinded allocation was guaranteed by storing the randomised list in an electronic file whose access was restricted to those in charge of recruitment. Participants were blinded as to which arm of the study group they belonged to. The data collectors were blinded to the outcome of the study as they did not have access to the results. During analysis, blinding was ensured by coding the intervention arm and the control arm. Those in charge of analysis did not know which group was intervention or control.

The intervention group had monthly sessions of Motivational Interviews with three trained physician research assistants who had undergone 20 hours of prior training in MI. They received intensive training in MI from basic principles to reflective listening, resolving ambivalence, working with resistance and change evocation. The training was conducted in the seminar room of the Department of Family Medicine, ISTH. It was conducted for them by professionals who had undergone similar training from professionals and had wide experience in the training and administration of MI. Training sessions were in the form of didactic lectures and practical sessions. Knowledge and skills of MI were assessed before and after the training sessions which lasted a total of 20 hours using a pre- and post-training questionnaire.

The MI sessions were at baseline, and then monthly for six months. This was in addition to the standard care given to patients with hypertension coming to the hospital. The control group had only the standard care without the Motivational Interview Session. This was administered by three physician research assistants who had not undergone prior training in MI. Outcome measurement was determined for both groups at six months. The primary outcome of the study was lifestyle modification at six months while the secondary outcome was blood pressure control at six months. The flow chart is illustrated in Fig 2.

**Fig 2:**
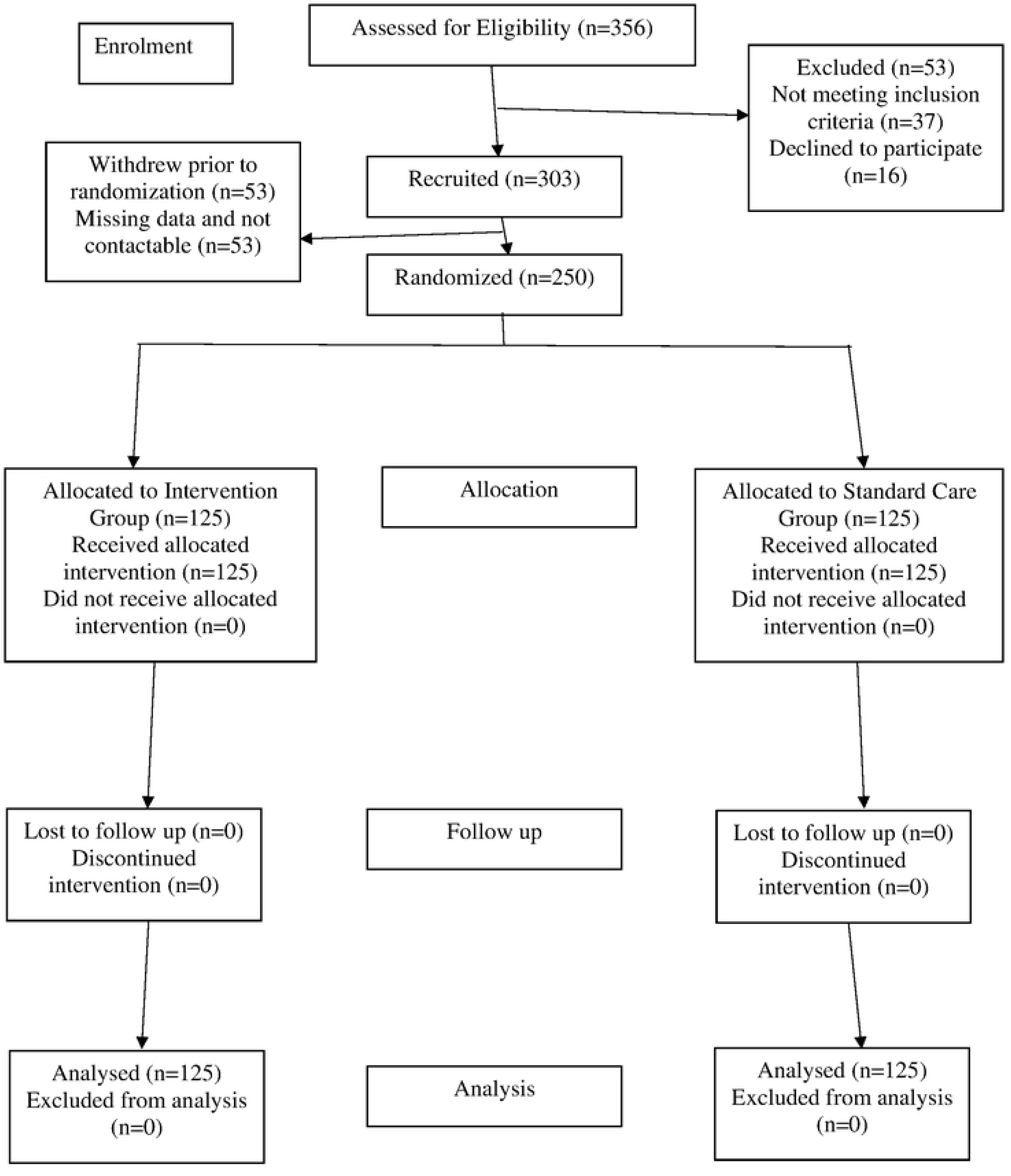
Consort Flow Diagram

### Study Instruments

A semi-structured questionnaire was employed for data collection. The questionnaire was made up of both open and close-ended questions covering relevant information on patients’ demographic information, employment status, and occupation. The questionnaire called Lifestyle Factors in Hypertension (LFH) Questionnaire was developed by the researcher in line with the study objectives. It was made up of four sections namely Sociodemographic data; Medical History; Lifestyle History; and Clinical Findings.

- Section A contained questions assessing participants’ sociodemographic characteristics such as age; sex; occupation which was categorised as civil servant, artisan, farmer, trader and unemployed (Housewives and students were categorised as unemployed); educational status, whether non-formal, primary, secondary or tertiary level of education; ethnic group; monthly income; and NHIA status.
- Section B assessed the medical history of respondents including the duration of hypertension, the current anti-hypertensive medications patients were taking, whether patients were regular on medication or not, and if not regular, the reasons for skipping medications.
- Section C contained questions on the lifestyle history of respondents. Physical activity, fruit and vegetable consumption, salt intake, smoking, and alcohol consumption were determined.
- Section D was the clinical findings from study participants.

### Validity of the Questionnaire

The questionnaire was evaluated by a team of experts in Family Medicine, Lifestyle Medicine, Community Medicine and Mental & Behavioural Health. It was also given to Cardiology, Epidemiology, Biostatistics, Exercise Medicine, and Nutrition Medicine experts. The questionnaire was also subjected to construct and content validity to ensure all areas of the study objectives were covered. Corrections and modifications were made where necessary.

### Reliability of the Questionnaire

To ensure the reliability and internal consistency of the questionnaire, it was administered to 22 patients with hypertension attending the Family Medicine Clinics of ISTH who met the study’s eligibility criteria. The data collected were coded, entered into SPSS statistical software and subjected to Cronbach’s Alpha coefficient. A Cronbach’s alpha coefficient of 0.85 was obtained. This was done before the commencement of the study.

### Variables and their Definitions/Measurements

#### Level of Physical Activity

Their level of physical activity was assessed. Those who met the WHO criteria of at least 30 minutes of daily moderate to intense physical activity for at least five days a week or a cumulative 150 minutes of moderate to intense physical activity per week were categorised as physically active while the others were categorised as inactive (sedentary). Moderate to intense physical activity as defined by the study was based on the WHO definition and included brisk walking, jogging, dancing, cycling, skipping, swimming and running.^44^

#### Salt intake

Their salt intake was assessed by asking if they added extra salt to food after serving. Those who added extra salt were categorised as taking excess salt while the others were categorised as taking moderate salt.^45^

#### Fruits and Vegetables intake

The number of servings of fruits and vegetables consumed daily was also assessed. A serving of fruit is a moderate-sized fruit while a serving of vegetables is half a cup of raw or cooked vegetables. Consumption of five or more servings of fruits and vegetables per day was considered adequate. Participants who consumed less than five servings were categorised as not having adequate intake of fruits and vegetables.^46^

#### Tobacco smoking

Tobacco smoking was assessed using Pack Years which was determined by multiplying the number of packets of cigarettes smoked per day by the duration of smoking in years. Participants were however categorised into smokers for those who currently smoked and non-smokers for those who never smoked or have quit smoking as no amount of tobacco smoking is safe.^47^

#### Alcohol Consumption

Alcohol consumption was determined and for those who drank alcohol, the number of units was assessed. A unit of alcohol was determined by multiplying the total volume in millilitres of drink consumed by the Alcohol by Volume (ABV) of the particular drink and dividing the result by 1,000. Patients were categorised as drinkers and non-drinkers as studies have shown that even a unit of alcohol increases the risk of hypertension and no level of alcohol is safe for consumption.^48,49^

#### Height (in meters)

It was taken using a portable stadiometer (Secca 240 wall mounted, Hamburg Germany) and recorded to the nearest 0.01 metre. With the patient barefooted and caps/headgear removed, the patient was made to stand erect with the back against the metre and the value was recorded.

#### Weight (in kilograms)

It was measured using a weighing scale (Secca 770 Floor Digital Scale, Hamburg Germany) with each subject in light clothing and bare feet. Weight was recorded to the nearest 0.1kg.

#### Body mass index (BMI)

It was determined by dividing the weight (in kilograms) by the square of the height (in meters). Patients were categorized based on their BMI as Underweight (<18kg/m^2^), normal weight (18 to 24 kg/m^2^), overweight (25 to 29 kg/m^2^), and obese (≥30 kg/m^2^).

#### Waist Circumference (in metres)

It was measured using a flexible, non-stretchable tape rule. It was measured horizontally at a point midway between the inferior costal margin and the iliac crest.

#### Hip Circumference (in metres)

It was also measured using the tape rule above at the level of the greater trochanter which is the widest diameter of the buttocks.

#### Waist Hip Ratio (WHR)

This was determined by dividing the waist circumference (in metres) by the hip circumference (in metres). Patients with WHR of < 1.0 for men and < 0.9 for women were categorized as normal while others were categorized as obese.^50^

#### Waist Height Ratio (WHtR)

It was determined by dividing the waist circumference (in metres) by the height (in metres). Patients were categorized as obese if their WHtR was ≥ 0.5 and not obese if their WHtR was < 0.5.^50^

#### Pulse Rate (beats per minute)

was obtained using an Omron digital sphygmomanometer (OMRON M2 Classic Intellisense)^®^ which measures both pulse and blood pressure accurately.

#### Office SBP and DBP

were obtained using an Omron digital sphygmomanometer (OMRON M2 Classic Intellisense)^®^ to measure BP correctly using the appropriate cuff sizes with an average of three BP measurements at 10 minutes intervals and adapted cuff (as shown in Table 1) at the brachial artery while the patient was seated in a chair with the back supported and the arm bare and at heart level. Blood pressure was measured in both arms and the arm with the higher BP value will be chosen. Three consecutive measurements were obtained; the first 10 minutes after sitting down, the second and the third after 10 minutes apart. The mean of the last two systolic values and the mean of the last two diastolic values of each subject were recorded as the office SBP and the office DBP respectively. A large adult-sized cuff was used to measure BP in individuals with large arms. Participants were told to avoid Tobacco, exercise and caffeine at least 30 minutes before BP measurements were taken. Respondents were categorised as normal if their blood pressure was ≤ 120/80mmHg; pre hypertensive if their blood pressure was between 121-139/81-89mmHg and hypertensive if their blood pressure was ≥ 140/90mmHg. This was based on the JNC 7 classification of hypertension.^51^

**Table 1:** Sociodemographic Characteristics of Respondents.

| Variable | Intervention<br>N=125<br>n (%) | Control<br>N=125<br>n (%) | Total<br>N=250<br>N (%) | $\chi^2$ | p-value |
| --- | --- | --- | --- | --- | --- |
| Age as at last birthday (years) |  |  |  |  |  |
| < 40 | 31 (24.8) | 15 (12.0) | 46 (18.4) | 7.0516 | 0.070 |
| 40 – 49 | 21 (16.8) | 27 (21.6) | 48 (19.2) |  |  |
| 50 – 59 | 44 (35.2) | 48 (38.4) | 92 (36.8) |  |  |
| ≥ 60 | 29 (23.2) | 35 (28.0) | 64 (25.6) |  |  |
| Sex |  |  |  |  |  |
| Female | 78 (62.4) | 78 (62.4) | 156 (62.4) | 0.000 | 1.000 |
| Male | 47 (37.6) | 47 (37.6) | 94 (37.6) |  |  |
| **Marital status** |  |  |  |  |  |
| Single | 5 (4.0) | 7 (5.6) | 12 (4.8) | 5.111 | 0.164 |
| Married | 73 (58.4) | 62 (49.6) | 135 (54.0) |  |  |
| Separated/Divorced | 17 (13.6) | 30 (24.0) | 47 (18.8) |  |  |
| Widowed | 30 (24.0) | 26 (20.8) | 56 (22.4) |  |  |
| **Occupation** |  |  |  |  |  |
| Artisan | 14 (11.2) | 18 (14.4) | 32 (12.8) | 8.482 | 0.075 |
| Civil servant | 53 (42.4) | 33 (26.4) | 86 (34.4) |  |  |
| Farmer | 10 (8.0) | 19 (15.2) | 29 (11.6) |  |  |
| Trader | 41 (32.8) | 46 (36.8) | 87 (34.8) |  |  |
| Unemployed | 7 (5.6) | 9 (7.2) | 16 (6.4) |  |  |
| **Average monthly income (₦)** |  |  |  |  |  |
| < 50,000 | 32 (25.6) | 40 (32.0) | 72 (28.8) | 1.731 | 0.641<sup>#</sup> |
| 50,000 – 99,999 | 60 (48.0) | 58 (46.4) | 118 (47.2) |  |  |
| 100,000 – 199,999 | 28 (22.4) | 24 (19.2) | 52 (20.8) |  |  |
| ≥ 200,000 | 5 (4.0) | 3 (2.4) | 8 (3.2) |  |  |
| **Religion** |  |  |  |  |  |
| Christianity | 70 (56.0) | 64 (51.2) | 134 (53.6) | 1.486 | 0.526<sup>#</sup> |
| Islam | 55 (44.0) | 60 (48.0) | 115 (46.0) |  |  |
| Traditional religion | 0 (0.0) | 1 (0.8) | 1 (0.4) |  |  |
| **Ethnic group** |  |  |  |  |  |
| Esan | 67 (53.6) | 58 (46.4) | 125 (50.0) | 6.386 | 0.215<sup>#</sup> |
| Afenmai | 51 (40.8) | 62 (49.6) | 113 (45.2) |  |  |
| Others (Yoruba, Hausa, Igbo and Kogi) | 7 (5.6) | 5 (4.0) | 12 (4.8) |  |  |
| **Health Insurance Coverage** |  |  |  |  |  |
| Enrolee | 30 (24.0) | 27 (21.6) | 57 (22.8) | 0.175 | 0.676 |
| Non-Enrolee | 95 (76.0) | 98 (78.4) | 193 (77.2) |  |  |

Mean age ± SD in years = 51.5 ± 10.0
<sup>#</sup>Fisher's exact test

### Data Collection

A semi-structured interviewer-administered questionnaire was used to obtain baseline information on sociodemographic characteristics, lifestyle patterns and clinical parameters from both groups. The intervention group were then given monthly sessions of MI counselling on the adoption of positive lifestyle modification in addition to standard care for hypertension for six months while the control group had standard care for hypertension which included routine counselling on lifestyle modification monthly after which outcome parameters were determined.

All participants (intervention arm and control arm) however received standard care for the management of hypertension which included routine history, examination, investigations, medications, and health education on healthy lifestyle choices.

#### Training of other Research Assistants

There was employment and training of research assistants, and pre-testing of questionnaires before the commencement of the fieldwork. They were trained for two days in the area of the job description. The research assistants were being monitored daily throughout the study. Completed questionnaires were kept to prevent unauthorized people from gaining access to them. Adequate mechanisms were put in place to safeguard and guarantee data accuracy, and quality and devoid of any bias, editing of completed questionnaires, and data entry. Such mechanisms included coding of the data, pass-wording of the computer containing the data to avoid third-party access, and the use of only one computer for entering the information.

### Data Analysis

Data were collected from both groups and analysed using Stata, version 17 (StataCorp LLC, College Station, Texas, USA). The background characteristics of respondents were analysed descriptively and reported with their proportions. The Shapiro-Wilk test was used to assess the normality of continuous variables while the McNemar chi-square test and the Paired t-test were used to compare participants’ baseline and characteristics. Lifestyle modification (physical activity, diet, alcohol reduction, and smoking cessation) was measured and McNemar’s chi-square test was used to compare participants’ outcome variables. The difference in BP from baseline measurement to the end of the study was calculated for each patient. The McNemar chi-square test was used to compare outcome variables while the paired t-test was used to compare the difference in mean BP between baseline BP and at 6 months for the control and intervention groups. Furthermore, blood pressures were categorized as either being controlled or not and presented as a proportion. Data was expressed as frequencies and percentages. Cohen’s d was applied in the assessment of the effect of MI. For all tests, a p-value < 0.05 was considered statistically significant.

### Ethical Approval

Ethical approval was sought and obtained for the study from the Ethics and Research Committee of Irrua Specialist Teaching Hospital, where the research was conducted (ISTH/HREC/20230802/446). Written consents were obtained from respondents before the study after a detailed explanation to them. Researchers and research assistants have undergone prior ethical training, especially on the consenting of participants. Research assistants obtained consent from patients before recruitment and randomization. The consent was translated into the local dialect with the aid of trained assistants who were proficient in the local dialect. Strict confidentiality was ensured and data were encrypted and protected from third parties.

### Trial Registration

This randomised clinical trial is registered with the Pan African Clinical Trial Registry (www.pactr.org) database. The unique identification number for the registry is PACTR202301917477205.

## RESULTS

A total of 356 hypertensive patients were screened for participation in the study, out of whom, 37 did not meet the eligibility criteria and 16 declined to participate, leaving 303 who met the eligibility criteria and agreed to participate (Fig 2). However, among the 303 agreed participants, 53 did not provide complete information at baseline and were thus excluded from the RCT, leaving 250 participants for inclusion in the study (effective response rate: 250/319 eligible participants=78.4%). All the 250 patients completed the study over the nine months (April to December 2023) and their results were analysed per protocol.

### Sociodemographic Variables

The age of the participants ranged from 24 to 65 years with a mean age of 51.5 ± 10.0 years. The majority of participants were in their 50s 92 (36.8 per cent) followed by those who were aged 60 and above 65 (26.0 per cent). Participants were mostly females 156 (62.4 per cent), married 135 (54.0 per cent), and Christians 134 (53.6 per cent). They were predominantly traders 87 (34.8 percent) and civil servants 86 (34.4 percent) with most participants having an average monthly income of between N50,000 and N99,999 118 (47.2 percent). Half of the participants were Esan by tribe from Edo Central Senatorial district where the study was conducted 125 (50.0 per cent) while 113 (45.2 per cent) were from Afenmai tribe in Edo North Senatorial District, the two districts mainly served by the hospital. The majority of patients did not have health insurance coverage. There was no significant difference between the sociodemographic characteristics of both the intervention and control groups. Table 1.

### Lifestyle Pattern of Respondents at Baseline

The lifestyle pattern of respondents at baseline is illustrated in Table 2. About three-quarters of respondents, 184 (73.6 per cent) were not physically active based on the WHO definition of 150 minutes of moderate to intense exercise weekly. The majority of the study participants took moderate salt 139 (55.6 per cent), did not smoke 210 (84.0 per cent), and did not take alcoholic drinks 193 (78.0 per cent). However, only 37 (14.8 per cent) participants consumed up to five servings of fruits and vegetables daily. There was no significant difference in the lifestyle pattern of both the intervention and control groups.

**Table 2:**
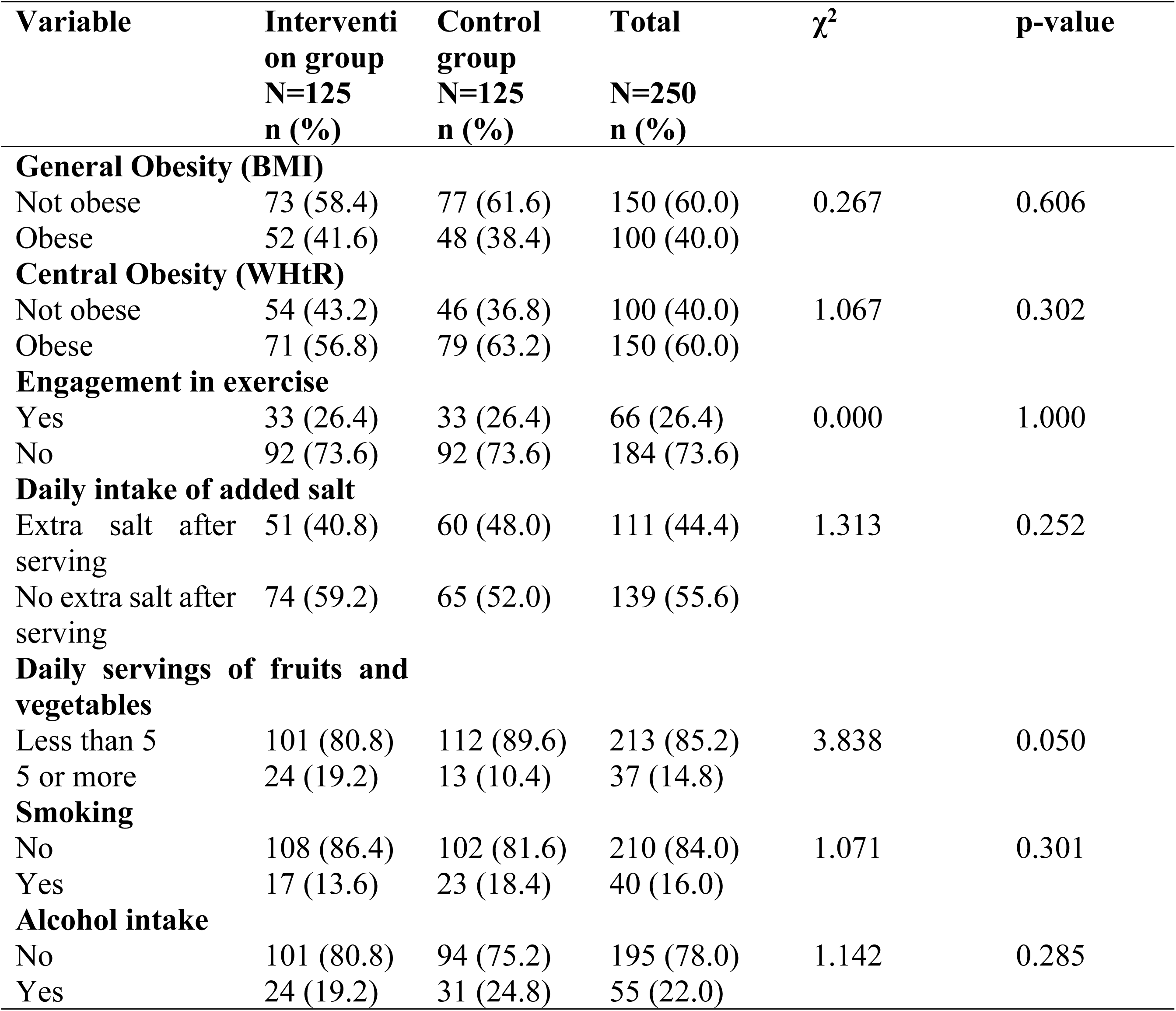
Lifestyle Pattern of Respondents at Baseline.

### Blood Pressure Control at Baseline

The majority of respondents 166 (66.4 per cent) had elevated systolic blood pressure at baseline, while 152 (60.8 per cent) had elevated diastolic blood pressure at baseline. The mean systolic blood pressure of patients was 144 ± 23 while their mean diastolic blood pressure was 89 ± 15. The blood pressure of respondents at baseline is shown in Table 3.

**Table 3:** Blood Pressure Measurements at Baseline.

| Variable | Frequency | Per cent |
| --- | --- | --- |

|  | (n=250) | (%) |
| --- | --- | --- |
| **Systolic blood pressure (mmHg)** |  |  |
| < 120 | 26 | 10.4 |
| 120 – 139 | 58 | 23.2 |
| ≥ 140 | 166 | 66.4 |
| **Diastolic blood pressure (mmHg)** |  |  |
| < 80 | 36 | 14.4 |
| 80 – 89 | 62 | 24.8 |
| ≥ 90 | 152 | 60.8 |

Mean ± SD of systolic blood pressure (mmHg) = 144 ± 23
Mean ± SD of diastolic blood pressure (mmHg) = 89 ± 15

The blood pressure control at baseline is illustrated in Fig 3. The blood pressure control rate was 24 percent with over three quarter of respondents having uncontrolled blood pressure at the commencement of the study.

**Fig 3:**
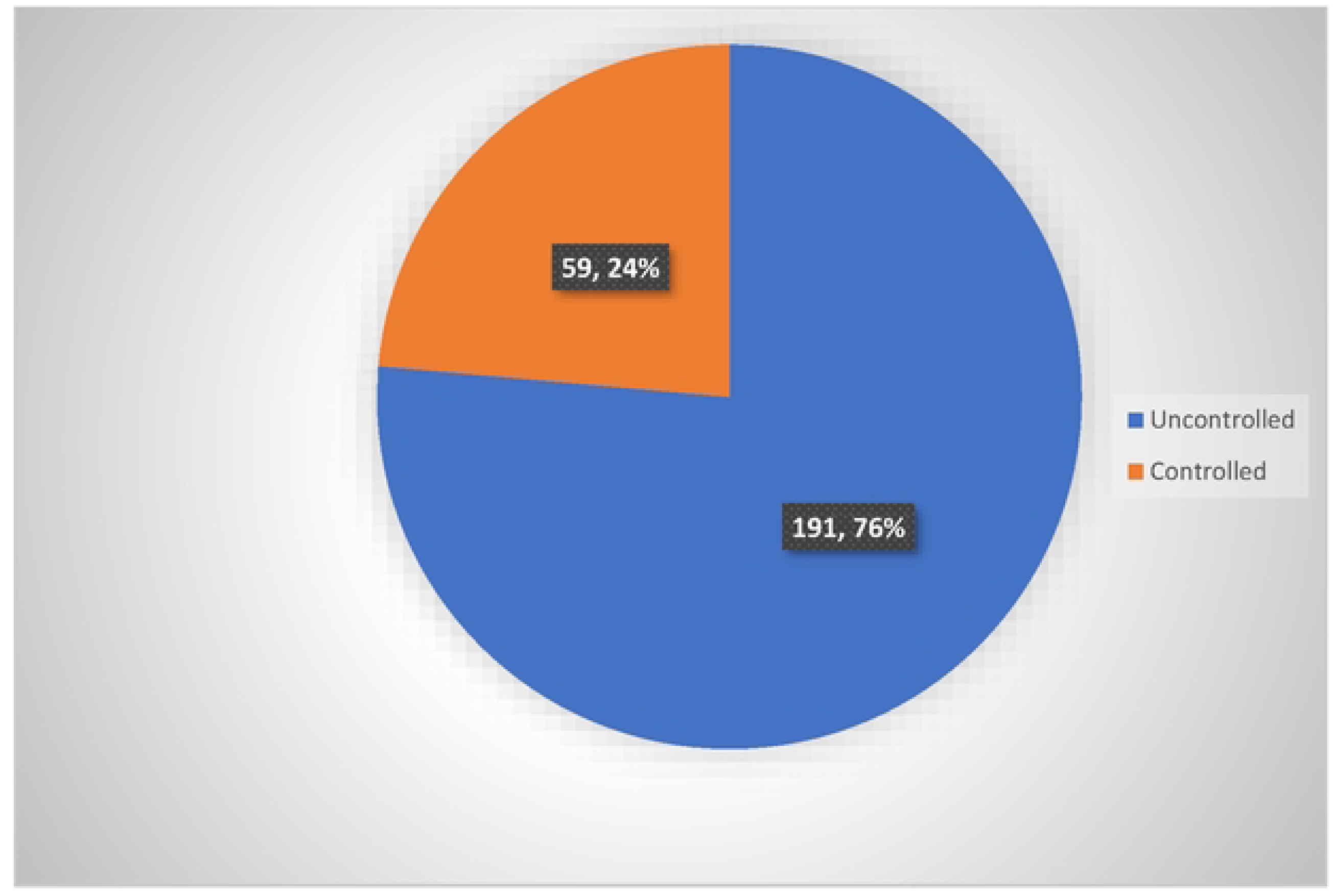
Blood Pressure control at Baseline (N=250)* *Controlled BP is systolic bp<140mmHG or diastolic bp <90mmHg while uncontrolled bp is systolic bp≥140mmHg or diastolic bp≥90mmHg

**Fig 4:**
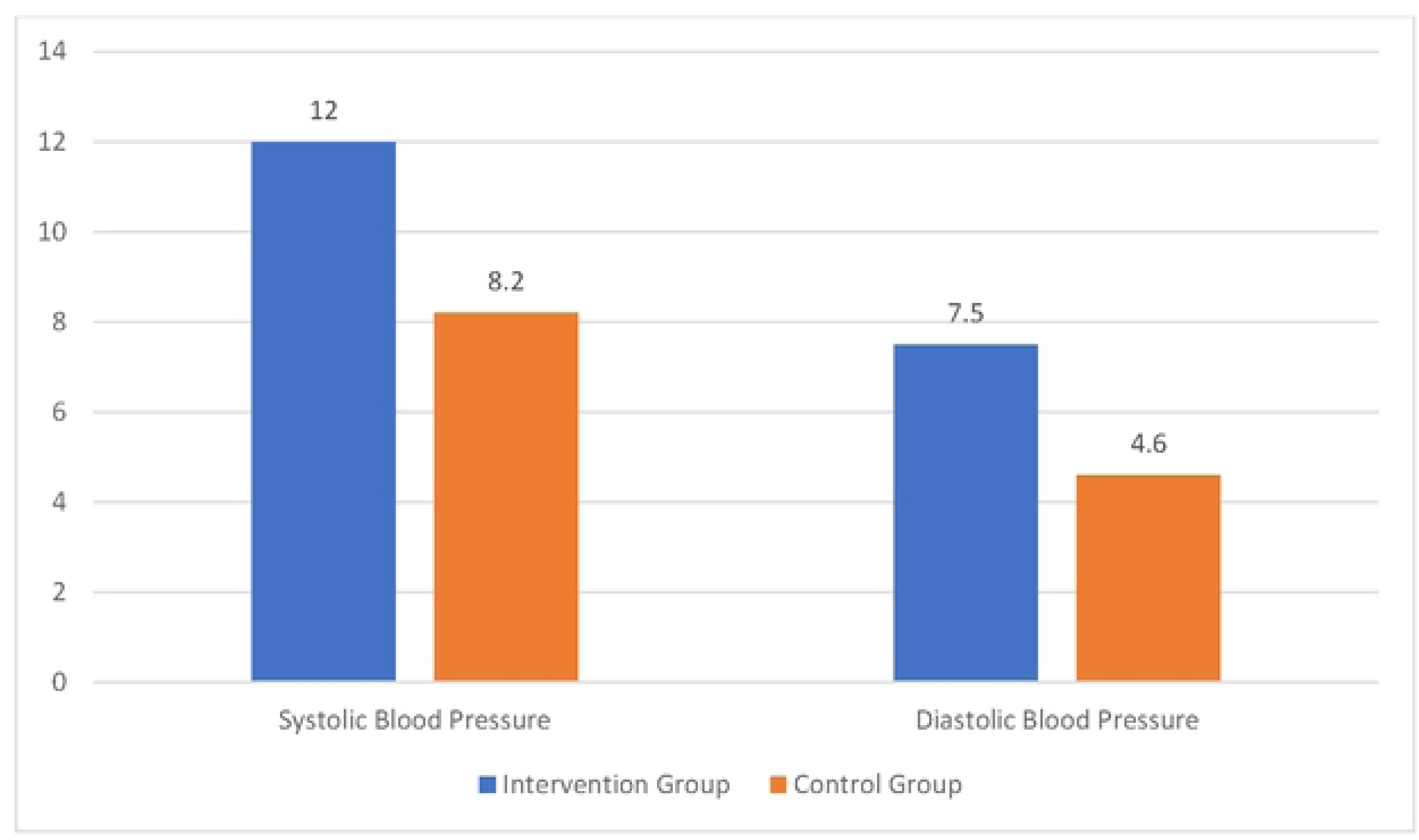
Mean Blood Pressure Reduction

### Effect of Motivational Interview on Lifestyle Modification

At the end of the six-month study, there was an improvement in the lifestyle pattern of the intervention group compared to control as shown in Table 4. This was statistically significant in all the parameters studied except smoking in which the difference was not statistically significant (p=0.150).

**Table 4:**
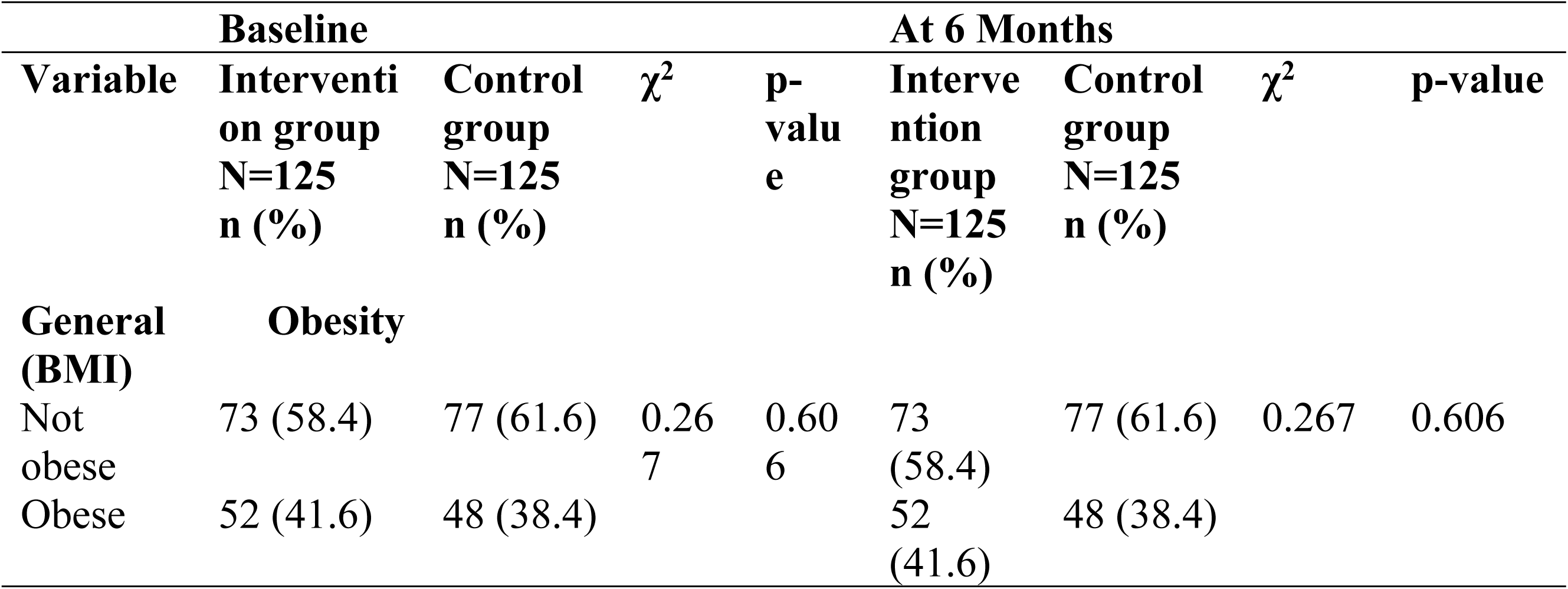
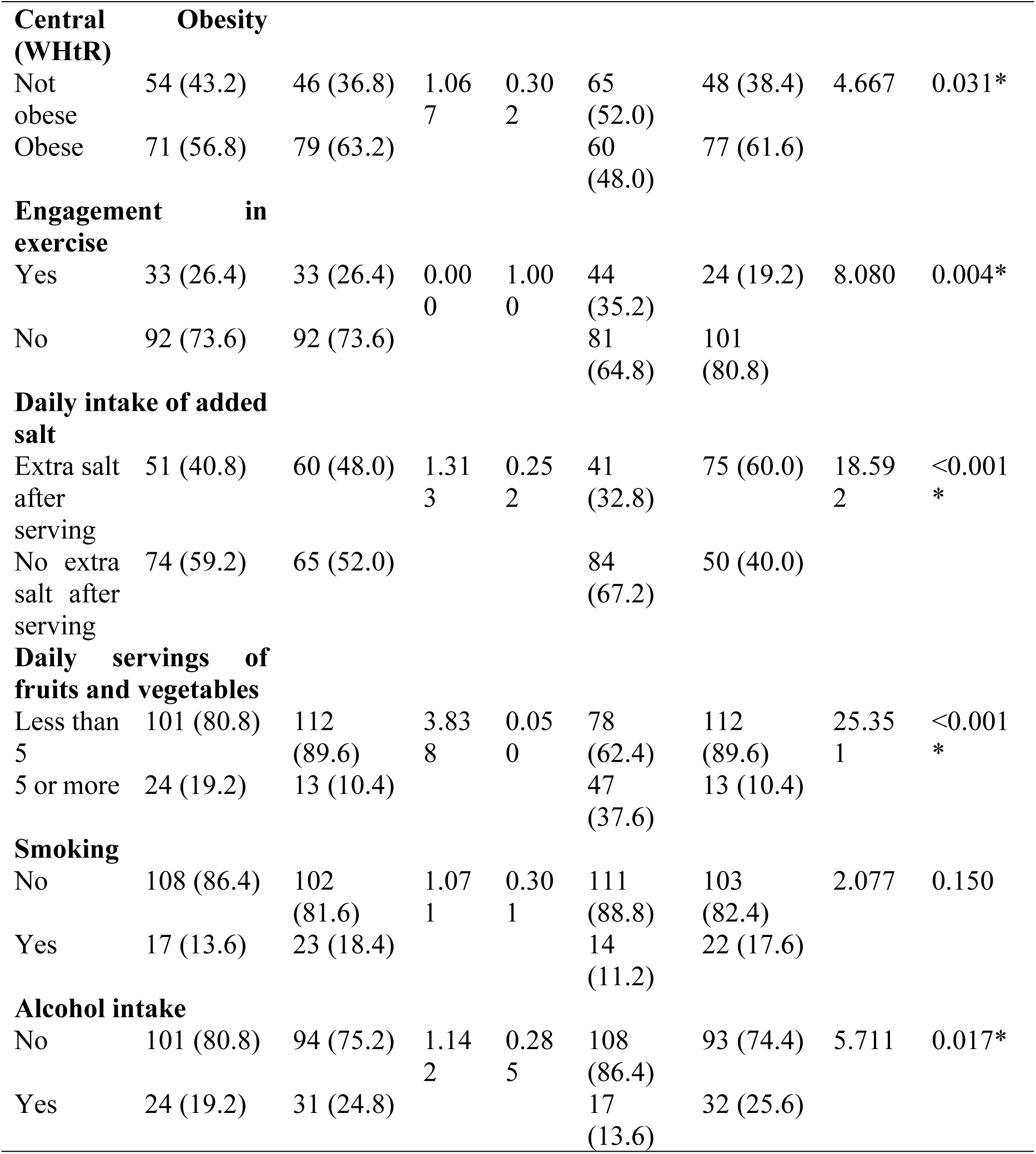
Effect of motivational Interview on Lifestyle Modification (N=250)

There was a statistically significant difference in the proportion of participants who engaged in exercise before and after the intervention (26.4 vs 35.2 per cent; p=0.004). Before the intervention 26.4 per cent of the intervention group engaged in exercise while after the intervention, the proportion significantly increased to 35.2 per cent.

A total of 23 participants changed their behaviour after the intervention, all of whom reported an increase in their fruit and vegetable intake from less than five servings to five or more servings. The difference was statistically significant (p<0.001).

Before the intervention, 40.8 per cent of respondents in the intervention group used extra salt after serving, while after the intervention, the percentage using extra salt dropped to 32.8 per cent. This difference was statistically significant (p<0.001).

Only three people reported a change in their smoking behaviour after the intervention, all of whom reported no longer smoking. Before the intervention, 13.6 percent of respondents in the intervention group smoked while after the intervention the proportion reduced to 11.2 percent. The difference was however not statistically significant (p=0.150).

A total of nine people reported a change in their alcohol intake after the intervention, of whom only one reported a change from not taking alcohol to now taking alcohol. All the other eight reported a change from taking alcohol to no longer taking alcohol. Before the intervention, 19.2 per cent of respondents in the intervention group reported using alcohol while after the intervention, the proportion using alcohol dropped to 13.6 per cent. The difference was statistically significant (p=0.017).

### Effect of Motivational Interview on Blood Pressure Control

Nearly half of those in the intervention group (48.0 per cent) had controlled blood pressure after the intervention compared to only about a third of those in the control group (32.8 per cent) who had controlled blood pressure. This association was statistically significant (p=0.014). This is illustrated in Table 5.

**Table 5:** Effect of Motivational Interview on Blood Pressure Control (N=250)

| **Variable** | **Control group**<br>**N=125** | **Intervention group**<br>**N=125** | **$\chi^2$** | **p-value** |
| --- | --- | --- | --- | --- |

|  | n (%) | n (%) |  |  |
| --- | --- | --- | --- | --- |
| **Baseline** |  |  |  |  |
| Uncontrolled | 96 (76.8) | 95 (76.0) | 0.022 | 0.882 |
| Controlled | 29 (23.2) | 30 (24.0) |  |  |
| **At 6 months** |  |  |  |  |
| Uncontrolled | 84 (67.2) | 65 (52.0) | 5.997 | 0.014* |
| Controlled | 41 (32.8) | 60 (48.0) |  |  |

\*Statistically significant

Post-intervention, the mean systolic and diastolic blood pressures reduced significantly by 12.0mmHg and 7.5mmHg respectively (p<0.001) as shown in Table 6. However, patients who had standard care only also had a significant mean systolic and diastolic blood pressure reduction of 8.2 and 4.6mmHg respectively (p<0.001).

**Table 6:** Effect of Motivational Interview on Blood Pressure Reduction.

| Variable | At baseline<br>Mean ± SD | After<br>intervention<br>Mean ± SD | Mean difference<br>(95% CI) | t<br>(paired t-test) | p-value |
| --- | --- | --- | --- | --- | --- |
| **Control Group** |  |  |  |  |  |
| Systolic BP in mmHg (n=125) | 145.3 ± 23.5 | 137.1 ± 16.6 | 8.2 (4.8 – 11.7) | 4.705 | <0.001 |
| Diastolic BP in mmHg (n=125) | 89.4 ± 15.4 | 84.8 ± 11.4 | 4.6 (2.4 – 6.8) | 4.184 | <0.001 |
| **Intervention Group** |  |  |  |  |  |
| Systolic BP in mmHg (n=125) | 141.9 ± 22.4 | 129.9 ± 14.6 | 12.0 (8.0 – 16.0) | 5.931 | <0.001 |
| Diastolic BP in mmHg (n=125) | 88.8 ± 14.1 | 81.3 ± 7.5 | 7.5 (5.2 – 9.7) | 6.573 | <0.001 |

## DISCUSSION

This Randomised Controlled Trial was aimed at determining the effect of motivational interviewing on lifestyle modification and blood pressure control among patients with hypertension attending the Family Medicine Clinics of Irrua Specialist Teaching Hospital, Irrua, Edo State, Nigeria.

### Effect of Motivational Interview on Lifestyle Modification among Respondents

This study demonstrated a significant improvement in lifestyle modification in the intervention group compared to control. Motivational interviewing was found to result in significant improvement in the WHtR (p=0.031), physical activity level (p=0.004), salt reduction (p<0.001), increased intake of fruits and vegetables (p<0.001), quality of sleep (p=0.002) as well as alcohol reduction (p=0.017). It also led to a reduction in BMI and smoking. However, the observed reduction did not reach statistical significance (BMI p=0.606; Smoking p=150).

Motivational interviewing led to a significant reduction in WHtR but not BMI. Whereas the World Health Organization has recommended BMI for the population-level measurement of obesity in adults as it does not vary with sex or age, it however cautions that it should be used as a rough guide since it may not measure the degree of fatness of the individuals.^52^ Measure of central obesity using WHtR has been recommended as the best measure of adiposity.^53^ Our study however found an improvement in both BMI and WHtR, this implies that MI if well delivered by trained health professionals can be useful in weight reduction programmes. This is important because obesity is a risk factor for non-communicable diseases such as CVDs (including stroke and heart disease), diabetes, osteoarthritis and other musculoskeletal disorders, and some cancers (such as breast, ovarian, colon, kidney and prostate cancers) with their risks increasing with increasing BMI.^52–54^ Obesity is also associated with premature death, hypertension, breathing difficulties, increased risk of fractures and other disability in adulthood when present in children.^52,54,55^ Findings from our study compare to findings from an RCT assessing the effect of MI on wellness which found a significant improvement in BMI and WC following intervention with MI over 6 months compared to controls.^38^ A prospective intervention study that involved the use of MI among hypertensive patients in Sweden over 1 year found a statistically significant improvement in weight, BMI and waist circumference.^56^

Our study revealed a notable correlation between motivational interviewing and engaging in physical activity. (p=0.004). This is in line with the findings of a systematic review on the effect of MI on lifestyle modification that found moderate evidence of the beneficial effect of MI on promoting physical activity participation.^34^ An intervention study using Telephone MI conducted among the Dutch population found a significant relationship between the percentage of MI adherent expressions and change in the level of PA from baseline to 47-week follow-up, with a small effect size (p<0.05).^37^

Participants, in our research, also raised their daily consumption of fruits and vegetables to 5 servings or more after undergoing motivational interviewing intervention. This increase was statistically significant (p<0.001). A similar intervention study found a statistically significant improvement in fruit consumption in patients following the use of MI compared to controls. However, the research did not identify a significant correlation between the utilization of motivational interviewing and vegetable intake.^37^

There was a significant reduction in salt intake (p=0.002) among participants in the intervention group after undergoing six monthly sessions of MI delivered by a physician. This is similar to the findings of an intervention study that used telehealth to deliver MI to patients which found a significant reduction in salt intake by 638 mg/day from a baseline of 2,919 mg/day (*P* < .001).^57^

We found a marginal improvement in the number of respondents who smoked following intervention from 13.6% at baseline to 11.2% at 6 months. However, the disparity was not statistically significant. (p=0.250). A systematic review revealed a noteworthy enhancement in smoking behaviours with motivational interviewing intervention as opposed to standard care.^31^

Yet another systematic review discovered moderate evidence supporting the positive impact of motivational interviewing on smoking cessation.^34^

A statistically significant decrease in alcohol consumption (p=0.039) was observed among participants in the intervention group. A systematic review found 8 out of 13 studies on MI providing consistent evidence that MI had a beneficial effect on the frequency and/or volume of alcohol consumption over a short period but with less consistent evidence after four months.^34^

### Effect of Motivational Interview on Blood Pressure Control among Respondents

In our study, we observed a noteworthy enhancement in blood pressure control within the intervention group, increasing from 24.0% at baseline to 48.0% at 6 months. In comparison, the control group started with a baseline blood pressure control rate of 23.2%, which rose to 32.8% at 6 months. This difference was statistically significant (p=0.014). Post-intervention, there was a mean reduction in systolic and diastolic blood pressures by 12.0mmHg and 7.5mmHg respectively and this was statistically significant (p<0.001). A systematic review on the effectiveness of MI on lifestyle modification in patients at risk of developing CVD or those with established CVD found MI to improve SDP and DBP but the difference was not statistically significant.^31^ An intervention study in Sweden found a significant reduction in SDP and DBP of 8.7mmHg and 4.0mmHg respectively after one year of lifestyle intervention involving the use of MI.^56^ MI through telephone counselling was also found to significantly reduce SDP and DBP at 12 months from baseline by 5.7 mm Hg and 4.1mmhg respectively.^57^ A systematic review and meta-analysis on the effect of motivational interview and blood pressure control found a statistically significantly greater reduction in systolic blood pressure (P = 0.001) in intervention programmes incorporating MI compared with no intervention or minimal intervention. The study also found programmes incorporating MI having statistically significantly greater reduction in systolic blood pressure (P = 0.040) compared to programmes with lower intensity interventions. The study also found a statistically significant benefit of diastolic blood pressure reduction from programmes incorporating motivational interviewing compared with no or minimal additional intervention (P < 0.001). Programmes incorporating motivational interviewing did not, however, outperform the lower-intensity interventions in diastolic blood pressure reduction (P = 0.600).^58^ The study found interventions combining individual and group delivery modes having a larger effect on diastolic blood pressure reduction compared to those delivered individually.

### Study Strengths

To the best of our knowledge, this study represents the initial randomized control trial conducted in Nigeria to assess the impact of motivational interviews on lifestyle modification and blood pressure control in individuals with hypertension.

The study examined the implementation of behavioural motivation to instigate lifestyle changes in adults with hypertension.

Additionally, the research investigated alternatives to pharmacotherapy by incorporating lifestyle modifications in the management of hypertension among adult individuals in Edo State, Nigeria.

### Study Limitations

The study was conducted in the Family Medicine Clinics of the hospital. Patients who accessed care in other clinics like the Cardiology and Nephrology clinics where a lot of patients with hypertension access care were left out. However, this would not significantly affect the findings from the study as most of the patients in these specialist clinics have other comorbidities.

Also, it was a hospital-based study, thus excluding hypertensive patients who sought care in other health facilities as well as non-health facilities.

Another limitation was that salt intake was accessed based on participants not adding extra salt to serving. This does not take into account participants who cook with excess salt or eat processed food with much salt.

Also, BP was measured in the clinic. An 8-day home measurement would have been more accurate.

## Conclusion

The study found that most patients with hypertension attending the Family Medicine Clinics of ISTH, Irrua, Edo State had central obesity and engaged in unhealthy lifestyle practices like living a sedentary lifestyle, and did not take adequate amounts of fruits and vegetables. They however mostly took moderate salt, did not smoke and did not consume alcohol. Their blood pressure was also poorly controlled.

The implementation of motivational interviewing resulted in a notable enhancement in physical activity, consumption of fruits and vegetables, reduction in salt intake, and diminished alcohol consumption. While there was progress in smoking cessation within the intervention group of patients compared to the control group, the observed difference did not reach statistical significance.

Likewise, the application of motivational interviewing resulted in a significant improvement in blood pressure control.

### Recommendations

Based on our findings, it is recommended to educate the general public about the risks associated with unhealthy lifestyles. Emphasis should be placed on promoting routine blood pressure checks and implementing measures to enhance blood pressure control in individuals with hypertension. Physicians are encouraged to address barriers to healthy living during consultations and prescribe appropriate lifestyle modifications.

Motivational interviewing, a technique beneficial for behaviour change, should be integrated into counselling sessions, conducted not only by physicians but also by nurses and social health workers. This approach applies to various health counselling aspects, including medication adherence. Extensive training in motivational interviewing for healthcare providers is advocated to enable effective implementation.

Further research on a larger scale is necessary to validate and apply these findings more broadly. Additionally, we recommend integrating motivational interviewing into the National Guidelines for Hypertension Management in Nigeria.

## Data Availability

Data used are available in the manuscript and also deposited in th Pan African Clinical Trial Registry (PACTR202301917477205)

## Acknowledgements

We thank the Head, Department of Family Medicine, Irrua Specialist Teaching Hospital, Irrua, Edo State, Nigeria, Dr BT Adewuyi and other staff of the department for making this study possible. We also thank the management of ISTH led by the Chief Medical Director, Prof Reuben Eifediyi for the support throughout the study. Finally, we acknowledge the Department of Family Medicine, University of Calabar Teaching Hospital for their immense contribution to the success of this project.

## Author Contributions

**Conceptualization**: Tijani Idris Ahmad OSENI, Afiong Oboko OKU, Roseline Ekanem DUKE, Ndifreke Ekpo UDONWA.

**Formal analysis**: Osahon OTAIGBE.

**Investigation**: Tijani Idris Ahmad OSENI, Sulaiman Dazumi AHMED, Osahon OTAIGBE.

**Methodology**: Tijani Idris Ahmad OSENI, Afiong Oboko OKU, Ifeoma N MONYE, Osahon OTAIGBE, Ndifreke Ekpo UDONWA.

**Supervision**: Afiong Oboko OKU, Roseline Ekanem DUKE, Ndifreke Ekpo UDONWA.

**Validation**: Tijani Idris Ahmad OSENI, Afiong Oboko OKU, Ifeoma N MONYE, Sulaiman Dazumi AHMED, Roseline Ekanem DUKE, Ndifreke Ekpo UDONWA.

**Writing** – **original draft**: Tijani Idris Ahmad OSENI, Afiong Oboko OKU, Ifeoma N MONYE, Sulaiman Dazumi AHMED, Roseline Ekanem DUKE, Osahon OTAIGBE, Ndifreke Ekpo UDONWA.

**Writing** – **review & editing**: Tijani Idris Ahmad OSENI, Afiong Oboko OKU, Ifeoma N MONYE, Sulaiman Dazumi AHMED, Roseline Ekanem DUKE, Osahon OTAIGBE, Ndifreke Ekpo UDONWA.

## REFERENCES

1. Adeloye D, Owolabi EO, Ojji DB, Auta A, Dewan MT, Olanrewaju TO, et al. Prevalence, awareness, treatment, and control of hypertension in Nigeria in 1995 and 2020: A systematic analysis of current evidence. J Clin Hypertens. 2021;23:963–977.

2. Schutte AE, Jafar TH, Poulter NR, Damasceno A, Khan NA, Nilsson PM et al. Addressing global disparities in blood pressure control: perspectives of the International Society of Hypertension. Cardiovascular Research. 2023; 119, 381–409 10.1093/cvr/cvac130

3. Gakidou E, Afshin A, Abajobir AA, Abate KH, Abbafati C, Abbas KM, Abd-Allah F, Abdulle AM, Abera SF, Aboyans V, Abu-Raddad LJ. Global, regional, and national comparative risk assessment of 84 behavioural, environmental and occupational, and metabolic risks or clusters of risks, 1990–2016: a systematic analysis for the Global Burden of Disease Study 2016. The Lancet. 2017 Sep 16;390(10100):1345–422.

4. Oguanobi NI. Management of hypertension in Nigeria: The barriers and challenges. J Cardiol Cardiovasc Med. 2021;6(1):23–25

5. WHO. Hypertension. Available at https://www.who.int/news-room/fact-sheets/detail/hypertension. Accessed on 12th October 2023

6. Adeke AS, Chori BS, Neupane D, Sharman JE, Odili AN. Socio-demographic and lifestyle factors associated with hypertension in Nigeria: results from a country-wide survey. Journal of Human Hypertension. 2022 Mar 24:1–6.

7. Murray CJ, Aravkin AY, Zheng P, Abbafati C, Abbas KM, Abbasi-Kangevari M, et al. Global burden of 87 risk factors in 204 countries and territories, 1990–2019: a systematic analysis for the Global Burden of Disease Study 2019. The lancet. 2020 Oct 17;396(10258):1223–1249.

8. Nwakile PC, Chori BS, Danladi B, Umar A, Okoye IC, Nwegbu M, et al. Removing the mask on hypertension (REMAH) study: Design; quality of blood pressure phenotypes and characteristics of the first 490 participants. Blood Pressure: 2019; 28:4, 258–267, DOI: 10.1080/08037051.2019.1612706

9. Idris A, Olaniyi O. Analysis of Economic Burden of Hypertension in Nigeria: Empirical Evidence from the Federal Capital Territory, Abuja. Nigerian Journal of Economic and Social Studies. 2020; 62(2): 147–174

10. Okoro TE, Edafe EA, Leader JT. The American Heart Association Classification of Blood Pressure and the Determinants of Hypertension among Medical Practitioners in Bayelsa State: A Cross-Sectional Study. J Biomed Res Clin Pract:2021;4(1):1–17.

11. Opara CJ, Maduka O. An Evaluation of Obesity and Hypertension among Primary School Teachers in an Urban Region of South-South Nigeria. J Hypertens Manag: 2020;6:050. 10.23937/2474-3690/1510050

12. Gnugesser E, Chwila C, Brenner S, Deckert A, Dambach P, Steinert JI et al. The economic burden of treating uncomplicated hypertension in Sub-Saharan Africa: a systematic literature review. BMC Public Health. 2022; 22:1507 10.1186/s12889-022-13877-4

13. Banigbe BF, Itanyi IU, Ofili EO, Ogidi AG, Patel D, Ezeanolue EE. High prevalence of undiagnosed hypertension among men in North Central Nigeria: Results from the Healthy Beginning Initiative. PLoS ONE: 2020; 15(11): e0242870. 10.1371/journal.pone.0242870

14. Odili AN, Chori BS, Danladi B, Nwakile PC, Okoye IC, Abdullahi U, et al. Prevalence, awareness, treatment and control of hypertension in Nigeria: data from a nationwide survey 2017. Global Heart. 2020;15(1).

15. Ojji DB, Baldridge AS, Orji IA, Shedul GL, Ojo TM, Ye J, et al. Characteristics, treatment, and control of hypertension in public primary healthcare centers in Nigeria: baseline results from the Hypertension Treatment in Nigeria Program. Journal of hypertension. 2022 May 21;40(5):888–96.

16. Chow CK, Gupta R. Blood pressure control: a challenge to global health systems. The Lancet. 2019 Aug 24;394(10199):613–5.

17. Zeng Z, Chen J, Xiao C, Chen W. A global view on prevalence of hypertension and human development index. Annals of Global Health. 2020;86(1): 67.

18. Princewel F, Cumber SN, Kimbi JA, Nkfusai CN, Keka EI, Viyoff VZ, et al. Prevalence and risk factors associated with hypertension among adults in a rural setting: the case of Ombe, Cameroon. The Pan African Medical Journal. 2019;34:147.

19. Noah SP, Etukumana EA, Udonwa N, Morgan UM. Risk Factors for Hypertension among Adult Patients Attending the General Outpatient Clinics of a Tertiary Hospital in Uyo, South-South Nigeria. West African Journal of Medicine. 2020;37(5):490–8.

20. Unger T, Borghi C, Charchar F, Khan NA, Poulter NR, Prabhakaran D, et al. 2020 International society of hypertension global hypertension practice guidelines. Hypertension 2020;75:1334–1357.

21. Carey RM, Wright JT Jr, Taler SJ, Whelton PK. Guideline-driven management of hypertension: an evidence-based update. Circ Res 2021;128:827–846.

22. Whelton PK, Carey RM, Aronow WS, Casey DE Jr, Collins KJ, Dennison Himmelfarb C, et al. 2017 ACC/AHA/AAPA/ABC/ACPM/AGS/APhA/ASH/ ASPC/NMA/PCNA guideline for the prevention, detection, evaluation, and management of high blood pressure in adults: a report of the American College of Cardiology/American Heart Association Task Force on Clinical Practice Guidelines. Circulation. 2018; 138:e484–e594.

23. Leung AA, Daskalopoulou SS, Dasgupta K, McBrien K, Butalia S, Zarnke KB, et al. Hypertension Canada’s 2017 guidelines for diagnosis, risk assessment, prevention, and treatment of hypertension in adults. Can J Cardiol. 2017;33:557–576.

24. Williams B, Mancia G, Spiering W, Agabiti Rosei E, Azizi M, Burnier M, et al. 2018 ESC/ESH guidelines for the management of arterial hypertension. Eur Heart J. 2018; 39:3021–3104

25. Challa HJ, Ameer MA, Uppaluri KR. DASH diet to stop hypertension. InStatPearls [Internet] 2021. StatPearls Publishing.

26. Andualem A, Gelaye H, Damtie Y. Adherence to Lifestyle Modifications and Associated Factors Among Adult Hypertensive Patients Attending Chronic Follow-Up Units of Dessie Referral Hospital, North East Ethiopia. Integrated Blood Pressure Control. 2020;13:145–156, DOI: 10.2147/IBPC.S275575

27. Elbur AI. Level of adherence to lifestyle changes and medications among male hypertensive patients in two hospitals in Taif; Kingdom of Saudi Arabia. Int J Pharm Sci. 2015;7(4):168–172.

28. Verma P, Srivastava M, Ratan K. Assessment of extent of lifestyle modification among diagnosed patients of hypertension attending tertiary care hospital. Int J Med Health Sci. 2015;4(2):196–201.

29. Okwuonu C, Emmanuel C, Ojimadu N. Perception and practice of lifestyle modification in the management of hypertension among hypertensives in south-east Nigeria. Int J Med Biomed Res. 2014;3 (2):121–131. doi:10.14194/ijmbr.3.2.8

30. Ademe S, Aga F, Gela D. Hypertension self-care practice and associated factors among patients in public health facilities of Dessie town, Ethiopia. BMC Health Serv Res. 2019;19(1):51. doi:10.1186/s12913-019-3880-0 1

31. Lee WWM, Choi KC, Yum RW, Yu DS, Chair SY. Effectiveness of motivational interviewing on lifestyle modification and health outcomes of clients at risk or diagnosed with cardiovascular diseases: a systematic review. Int J Nurs Stud. 2016;53:331–341.

32. Nijs J, Wijma AJ, Willaert W, Huysmans E, Mintken P, Smeets R, et al Integrating motivational interviewing in pain neuroscience education for people with chronic pain: a practical guide for clinicians. Physical therapy. 2020;100(5):846–59.

33. Zabolypour S, Alishapour M, Behnammoghadam M, Larki RA, Zoladl M. A Comparison of the Effects of Teach-Back and Motivational Interviewing on the Adherence to Medical Regimen in Patients with Hypertension. Patient Preference and Adherence.2020;14:401–410.

34. Sawyer AT, McManus K. Understanding patient experiences in a motivational interviewing intervention to improve whole-person lifestyle among individuals with hypertension or type 2 diabetes: a qualitative focus group study. International journal of qualitative studies on health and well-being. 2021 Jan 1;16(1):1978373.

35. Frost H, Campbell P, Maxwell M, O’Carroll RE, Dombrowski SU, Williams B, et al. Effectiveness of motivational interviewing on adult behaviour change in health and social care settings: a systematic review of reviews. PloS one. 2018 Oct 18;13(10):e0204890.

36. Miller WR, Rollnick S. Motivational Interviewing, Helping People Change. New York: The Guilford Press; 2013

37. Mesters I, van Keulen HM, de Vries H, Brug J. Intervention fidelity of telephone motivational interviewing on physical activity, fruit intake, and vegetable consumption in Dutch outpatients with and without hypertension. International Journal of Behavioral Medicine. 2023 Feb;30(1):108–121.

38. Sawyer AT, Wheeler J, Jennelle P, Pepe J, Robinson PS. A randomized controlled trial of a motivational interviewing intervention to improve whole-person lifestyle. Journal of Primary Care & Community Health. 2020 May;11:2150132720922714.

39. Del Rio Szupszynski KP, de Ávila AC. The Transtheoretical Model of Behavior Change: Prochaska and DiClemente’s Model. Psychology of Substance Abuse: Psychotherapy, Clinical Management and Social Intervention. 2021:205–216.

40. Oseni TIA, Oku AO, Udonwa NE, Duke RE. Effect of Motivational Interviewing on Lifestyle Modification among Patients with Hypertension Attending the Family Medicine Clinics of ISTH, Irrua, Nigeria (Milmaph Study) – A Randomised Control Trial Study Protocol. West African Journal of Medicine. 2024; 41(2): 126–134

41. Irrua Specialist Teaching Hospital. Five – year strategic plan (2007-2011). 2022. https://fdocuments.in/doc.

42. Walters SJ, Jacques RM, dos Anjos Henriques-Cadby IB, Candlish J, Totton N, Xian MT. Sample size estimation for randomised controlled trials with repeated assessment of patient-reported outcomes: what correlation between baseline and follow-up outcomes should we assume?. Trials. 2019 Dec;20:1–6.

43. Steffen PL, Mendonça CS, Meyer E, Faustino-Silva DD. Motivational interviewing in the management of type 2 diabetes mellitus and arterial hypertension in primary health care: an RCT. American Journal of Preventive Medicine. 2021 May 1;60(5):e203–12.

44. WHO: WHO Guidelines on physical activity and sedentary behaviour. 2020. Available from https://www.who.int/publications/i/item/9789240015128 (Accessed on 2nd November, 2023).

45. Odili AN, Chori BS, Danladi PC, Nwakile PC, Ogedengbe JO, Nwegbu MMA, et al. Salt intake in Nigeria: a nationwide population survey. European Health Journal. 2020;22(12):2266–75.

46. Locke A, Schneiderhan J, Zick SM. Diets for health: Goals and guidelines. American Family Physician. 2018; 97: 721–728.

47. Masters NJ. Smoking pack years calculator. British Journal of General Practice. 2020; 70: 230.

48. WHO. No level of alcohol consumption is safe for our health. 2023. Available from https://www.who.int/europe/news/item/04-01-2023-no-level-of-alcohol-consumption-is-safe-for-our-health (Accessed on 2nd November, 2023).

49. Chi FW, Parthasarathy S, Palzes VA, Kline-Simon A H, Weisner CM, Satre DD, et al. Associations between alcohol brief intervention in primary care and drinking and health outcomes in adults with hypertension and type 2 diabetes: a population-based observational study. BMJ open, 2023;13(1), e064088.

50. Alwadeai KS, Almeshari MA, Alghamdi AS, Alshehri AM, Alsaif SS, Al-Heizan MO, et al. Relationship Between Heart Disease and Obesity Indicators Among Adults: A Secondary Data Analysis. Cureus. 2023 Mar 27;15(3).

51. Rahman M, Halder HR, Yadav UN, Mistry SK. Prevalence of and factors associated with hypertension according to JNC 7 and ACC/AHA 2017 guidelines in Bangladesh. Scientific reports, 2021;11(1):1–10.

52. WHO. Obesity and overweight. 2021. Available at: https://www.who.int/news-room/fact-sheets/detail/obesity-and-overweight (Accessed 10^th^ November, 2023).

53. Moosaie F, Fatemi Abhari SM, Deravi N, Karimi Behnagh A, Esteghamati S, Dehghani Firouzabadi F, et al. Waist-to-height ratio is a more accurate tool for predicting hypertension than waist-to-hip circumference and BMI in patients with type 2 diabetes: A prospective study. Frontiers in Public Health. 2021 Oct 7;9:726288.

54. Ayogu RN, Ezeh MG, Udenta EA. Epidemiological characteristics of different patterns of obesity among adults in rural communities of south-east Nigeria: a population-based cross-sectional study. BMC nutrition, 2022;8(1):1–10.

55. Chukwuonye II, Ohagwu KA, Ogah OS, John C, Oviasu E, Anyabolu EN, et al. Prevalence of overweight and obesity in Nigeria: systematic review and meta-analysis of population-based studies. PLOS Global Public Health. 2022 Jun 10;2(6):e0000515.

56. Lönnberg L, Ekblom-Bak E, Damberg M. Reduced 10-year risk of developing cardiovascular disease after participating in a lifestyle programme in primary care. Upsala journal of medical sciences. 2020 Jul 2;125(3):250–256.

57. Schrauben SJ, Inamdar A, Yule C, Kwiecien S, Krekel C, Collins C, et al. Effects of dietary app-supported tele-counseling on sodium intake, diet quality, and blood pressure in patients with diabetes and kidney disease. Journal of Renal Nutrition. 2022 Jan 1;32(1):39–50.

58. Huang X, Xu N, Wang Y, Sun Y, Guo A. The effects of motivational interviewing on hypertension management: a systematic review and meta-analysis. Patient Education and Counseling. 2023 Apr 14:107760.

